# Brain growth and neurodevelopment after surgical treatment of infant post-infectious hydrocephalus in sub-Saharan Africa

**DOI:** 10.1101/2020.11.10.20229328

**Authors:** Steven J. Schiff, Abhaya V. Kulkarni, Edith Mbabazi Kabachelor, John Mugamba, Peter Ssenyonga, Ruth Donnelly, Jody Levenbach, Vishal Monga, Mallory Peterson, Venkateswararao Cherukuri, Benjamin C. Warf

## Abstract

**Importance:** Post-infectious hydrocephalus in infants is a major public health burden in sub-Saharan Africa.

**Objective:** To determine long-term brain growth and cognitive outcome after surgical treatment of infant post-infectious hydrocephalus in Uganda.

**Design:** Prospective follow-up of a previously randomized cohort.

**Setting:** Single center in Mbale, Uganda.

**Participants:** Infants (<180 days old) with post-infectious hydrocephalus.

**Interventions:** Endoscopic or shunt surgery.

**Main outcomes:** Bayley Scales of Infant Development (BSID-3) and brain volume on computed tomography (raw and normalized for age and sex) at 2 years after treatment.

**Results:** Eighty-nine infants were assessed for 2-year outcome. There were no significant differences between the two surgical treatment arms, so they were analyzed together. Raw brain volumes increased between baseline and 24 months (median change=361 cc, IQR=293 to 443, p<0.001), but almost all of this increase was seen in the first year (median change=381 cc, IQR=310 to 442, p<0.001), with very little change between 12 and 24 months (median change=-5 cc, IQR=-52 to 42, p=0.66). The fraction of those with a normal brain volume increased from 15% at baseline to 50% at 1 year, but then declined to 18% at 2 years. Substantial normalized brain volume loss was seen in 21% between baseline and year 2 and in 77% between years 1 and 2. The extent of brain growth in the first year was not associated with extent of brain volume changes in the second year. There were significant positive correlations between 2-year brain volume and all BSID-3 scores and BSID-3 changes from baseline.

**Conclusions and Relevance:** In sub-Saharan Africa, even after successful surgical treatment of infant post-infectious hydrocephalus, post-treatment brain growth stagnates in the second year. While the reasons for this are unclear, this emphasizes the importance of primary infection prevention strategies along with optimizing the child’s environment to maximize brain growth potential.

**Trial Registration:** ClinicalTrials.gov number, NCT01936272

**KEY POINTS:** *Question:* What is the brain growth and cognitive trajectory of infants treated for post-infectious hydrocephalus in Uganda?

*Findings:* In this prospective follow-up of a cohort of 89 infants, early normalization of brain volume after treatment was followed by brain growth stagnation in the second year, with many falling back into the sub-normal range. Poor brain growth was associated with poor cognitive outcome.

*Meaning:* Successful surgical treatment of hydrocephalus is not sufficient to allow for adequate brain growth and cognitive development. Interventions aimed at primary infection prevention and reducing comorbidities are needed to improve brain growth potential.

## INTRODUCTION

Infant hydrocephalus is common worldwide and especially so in many low-income countries, with an estimated incidence in sub-Saharan Africa alone of approximately 180,000 infants per year.^1^ A dominant contributor appears to be post-infectious hydrocephalus following neonatal sepsis.^2,3^ Surgical treatment of hydrocephalus in this setting has proven to be safe^4^ and with a favorable benefit-cost ratio.^5^ A recent US National Institutes of Health-funded randomized trial in Uganda has shown that the two available treatment options – ventriculoperitoneal shunt (VPS)^6,7^ and the newer endoscopic third ventriculostomy combined with choroid plexus cauterization (ETV/CPC)^8–13^ – are both able to relieve symptoms and allow the brain to grow at 1 year after treatment, often regaining normal size.^14^ These results also showed, for the first time, the importance of brain growth in infant hydrocephalus because of its positive correlation with cognitive outcome. This represents a paradigm shift in our thinking of hydrocephalus treatment, which has traditionally been aimed at reducing ventricle size and intracranial cerebrospinal fluid (CSF) volume; the randomized trial showed no such correlation between CSF volume and cognitive outcome.^14^ These early results from the trial, however, led to an important question: does successful surgical treatment in early infancy lead to sustained, long-term brain growth and improved cognitive outcome beyond the first year? That is, is this just a transient phenomenon that later stabilizes or even regresses, despite treatment? This is especially relevant since the exact cause and mechanism of the early brain growth after hydrocephalus treatment is not known and it is not until 2 to 3 years of age that the human brain achieves 80 - 90% % of its adult volume.^15^. The answers might have relevance to not just post-infectious hydrocephalus in sub-Saharan Africa, but also infant hydrocephalus of other causes worldwide. We have, therefore, continued to follow this cohort of randomized infants to determine long-term brain growth trajectories and correlations with developmental outcome 2 years after treatment, which we present herein.

## METHODS

### Study design and participants

Prospective follow-up was done of a cohort previously randomiezed to ETV/CPC or VPS for post-infectious hydrocephalus. This was conducted at the CURE Children’s Hospital of Uganda (CCHU), a freestanding pediatric neurosurgical hospital in Mbale, Uganda. Trial design and 1-year results have previously been published.^14^ In summary, eligible infants were <180 days old, meeting established criteria for post-infectious hydrocephalus,^3^ excluding those with active clinical CSF infection or CT evidence of congenital brain anomaly, severe anatomical brain distortion, or multi-loculated hydrocephalus. Written consent was obtained from the mother in her home language.

### Interventions

The ETV/CPC arm underwent a standard unilateral frontal approach with flexible endoscopy.^8^ If the floor of the third ventricle could not be opened or if there was intra-operative evidence of substantial prepontine cistern scarring (which correlates with a high rate of ETV/CPC failure) ^13^, a VPS was inserted instead.^16^ The VPS arm underwent placement of a Chhabra (Shahjahanpur, India) VPS as per usual CCHU protocol.^6,7^ Successful treatment was determined by previously described clinical and radiographic criteria.^3,6,8,17^ In the case of treatment failure, the treating surgeon determined the type of repeat operation performed (reopening of ETV, VPS placement or VPS revision).^18^

### Follow-up

Patients were seen for clinical assessment and head CT at approximately 12 months and 24 months after surgery. Home visits were conducted for those who failed to return for follow-up.

### Outcomes

Developmental outcome was measured by trained evaluators, who, under the supervision of a neuropsychologist, used a culturally modified Bayley Scales of Infant Development (BSID-3) to evaluate all children.^19^ Children wore hooded jackets to blind the evaluators to their treatment allocation. The Cognitive, Gross Motor, and Fine Motor scaled scores at 24 months after surgery were obtained. BSID-3 scaled scores range from 1 to 19, with a general population mean of 10 and standard deviation (SD) of 3. The language scale portion of the BSID was not administered, since at 24 months of age the complexity of language and the cultural irrelevance of the test could become confounding factors and we had no other validated instrument for language assessment at 24 months in any of the many local languages. Therefore, only the BSID-3 Cognitive and Motor Scales were administered. Baseline BSID-3 was performed within 1 week before surgery to one day after surgery and included Cognitive, Motor, and Language Scales.

### Brain and CSF Volumes

We employed previously described^14^ semi-automated image segmentation of each CT scan to calculate brain and CSF volume. Using normalized brain growth curves derived from 1,067 normal children’s normal MRI scans in the North American NIH Pediatric MRI Data Repository (http://pediatricmri.nih.gov/nihpd/info/index.html), the normative growth percentile for the age and sex of each patient was quantified.^20^ Normative data for Ugandan infants is not available. The individuals performing these volumetric assessments were blinded to the patients’ BSID-3 results.

### Statistical analyses

Comparison between treatment arms as an intention-to-treat analysis showed no significant differences in important outcomes, including 24-month BSID-3 scores and brain volume. Therefore, the treatment groups were combined for further analyses.

Brain volumes were converted to z-scores, based on age- and sex-corrected volume distributions for North American infants.^20^ We determined the proportion of patients who achieved *substantial brain growth* (an increase in brain volume z-score of 1 or more between baseline and 24 months), *substantial brain volume loss* (a decrease in brain volume z-score of 1 or more between baseline and 24 months), and *normal brain volume* (brain volume larger than 1 SD below the age- and sex-corrected mean). Paired data were compared with Wilcoxon Signed Rank test.

Analyses were performed to determine the relationship between brain volume and developmental outcome. Spearman correlation was assessed between all 24-month BSID-3 scores and brain and CSF volumes separately. Two-year BSID-3 scores and changes in BSID-3 scores were compared between those who did and did not achieve *substantial brain growth* and *substantial brain volume loss* using the Mann-Whitney test.

We used the Hodges-Lehmann estimator to determine differences between medians with confidence intervals.

The **Appendix** lists trial structure and personnel. The study received ethics board approval at CCHU, Boston Children’s Hospital, The Hospital for Sick Children, and The Pennsylvania State University. Funding was provided by National Institutes of Health via the Eunice Kennedy Shriver National Institute of Child Health & Human Development and the Fogarty International Center (project numbers R21TW009612 and R01HD085853), and additional support for image analysis and construction of normative brain growth curves from an NIH Director’s Transformative Award (project number R01AI145057). The trial was registered at Clinicaltrials.gov (NCT01936272).

## RESULTS

Between May 2013 and April 2015, 158 patients were screened (**Figure 1**), of whom 58 were excluded, leaving 100 eligible randomized patients (51 randomized to ETV/CPC and 49 to VPS). We stopped recruitment after randomization of the pre-determined number of eligible patients. Previously published primary analysis failed to show a difference in the trial’s primary outcome (12-month BSID-3 Cognitive Scaled Score).^14^ Baseline data for the cohort are shown in **Table 1**.

**Table 1.**
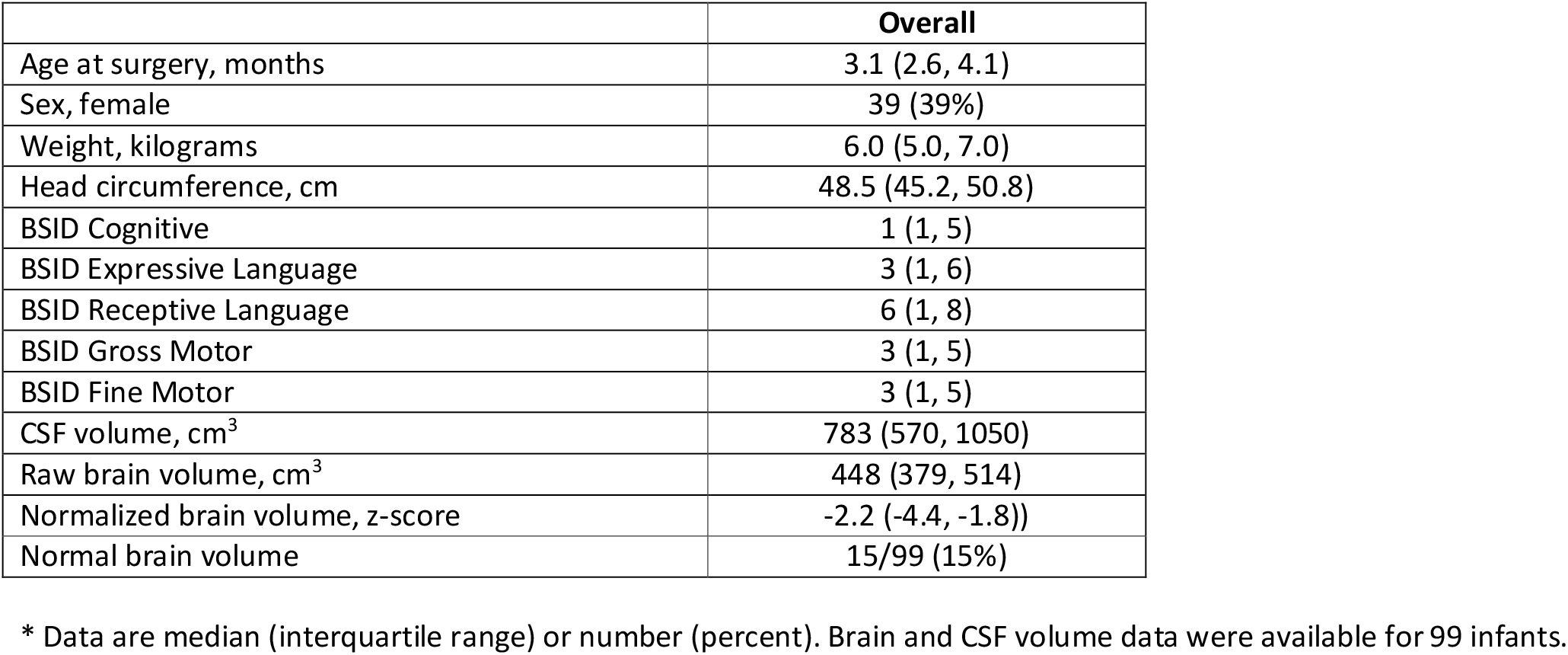
Patient characteristics at baseline*.

**Figure 1.**
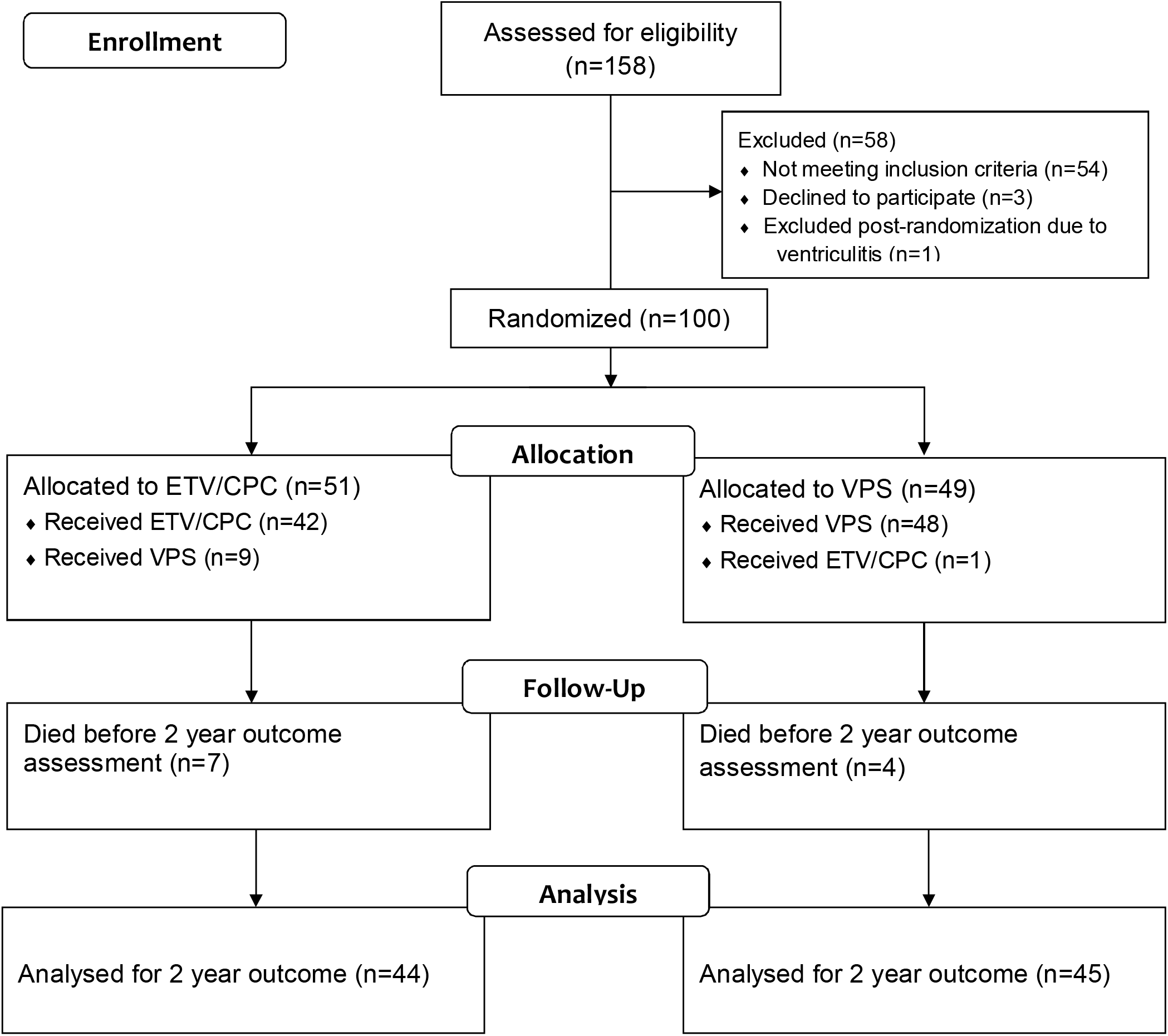
Enrollment, randomization, treatment, and follow-up.

### Treatment complications

There were 15 deaths (9 at <24 months): 1 attributed to treatment failure 8 months after ETV/CPC; 1 from infection at 5.5 months after initial ETV/CPC and then crossing over to VPS; 4 acute gastroenteritis (at 7, 12, 24, 29 months); 4 malnutrition (at 12, 12, 21, 30 months); 3 febrile illness (32, 33, 35 months); 1 pneumonia (at 22 months); 1 measles (at 24 months). All-cause mortality of 15% was comparable to our prior retrospective study of survival after treatment for post-infectious hydrocephalus in this population that found a 2-year mortality rate of approximately 20%.^21^

### Developmental outcome

Two-year developmental outcome was available for 89 patients (**Figure 1**). Between baseline and 2 years, only the BSID-3 Fine Motor score showed a significant improvement (p=0.004), with no significant changes for Cognitive or Gross Motor scores (p=0.18 and p=0.63, respectively) (**Table 2**).

**Table 2.** Outcome results at 24 months*.

|  | Overall |
| --- | --- |
| Head circumference, cm | 49.5 (46.9, 51.5) |
| Treatment failure | 33/100 (33%) |
| Infection | 4 (4.0%) |
| All-cause mortality | 15 (15%) |
| BSID Scaled Scores |  |
| BSID Cognitive | 4 (1, 5) |
| BSID Gross Motor | 1 (1, 5) |
| BSID Fine Motor | 5 (1, 8) |
| Change in BSID Scaled Scores from baseline |  |
| BSID Cognitive, change from baseline | 0 (-2, 3) |
| BSID Gross Motor, change from baseline | 0 (-3, 1) |
| BSID Fine Motor, change from baseline | 2 (-2, 4) |
| Brain volumes (raw) |  |
| Brain volume, baseline, cm<sup>3</sup> | 448 (379, 514) |
| Brain volume, 12 months, cm<sup>3</sup> | 813 (709, 941) |
| Brain volume, 24 months, cm<sup>3</sup> | 831 (725, 923) |
| Brain volumes (normalized for age and sex) |  |
| Normalized brain volume, 24 months, z-score | -2.6 (-3.9, -1.4) |
| Normal brain volume, 24 months, N (%) | 16/90 (18%) |
| Substantial brain growth from baseline to 24 months, N (%) | 23/89 (26%) |
| Substantial brain volume loss from baseline to 24 months, N (%) | 19/89 (21%) |
| Substantial brain volume loss from 12 to 24 months, N (%) | 69/90 (77%) |
| Substantial brain growth from 12 to 24 months, N (%) | 2/90 (2%) |

\* Data are median (interquartile range) or number (percent).

### Brain and CSF volume outcome

Baseline volume data were available for 99 patients (**Table 1**) and 24-month volume data for 89 (**Figure 2** and **Table 2)**. Raw brain volumes increased between baseline and 24 months (median change 361 cc, IQR 293 to 443, p<0.001), but almost all of this increase was seen in the first year (median change 381 cc, IQR 310 to 442, p<0.001), with very little change between 12 and 24 months (median change -5 cc, IQR -52 to 42, p=0.66, **Figure 2A and 2B**). Twenty-three patients (26%) demonstrated *substantial normalized brain growth* between baseline and 24 months, but these were almost all infants who had demonstrated that substantial growth in the first 12 months alone. Only 2 (2%) infants demonstrated substantial growth between 12 and 24 months. Normal brain volume was observed in 15% at baseline, 50% at 12 months, and only 18% at 24 months (**Figure 2C**). A minority of patients (21%) suffered *substantial normalized brain volume loss* at 24 months compared to baseline; but, 77% of all patients suffered substantial z score decline between 12 and 24 months (**Figure 2C**). There was no significant difference in the change in normalized brain volume in the second year between those who experienced *substantial normalized brain growth* in the first year (N=24, mean z score change -1.53) and those who did not (N=65, mean z score change -1.35, p=0.3)

**Figure 2.**
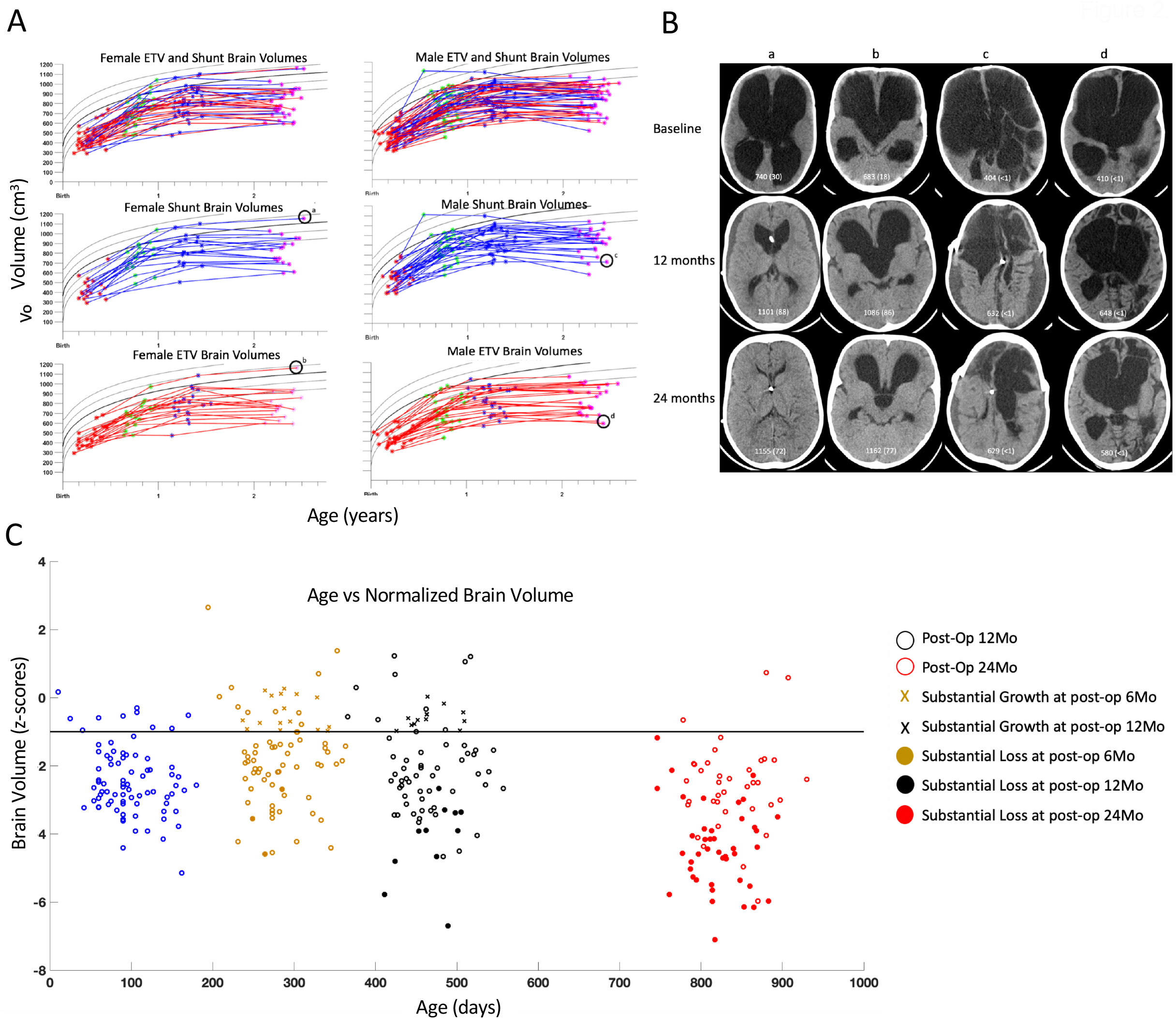
Brain volumes and imaging. In A are shown the raw brain volumes, in cm^3^, stratified by males, females, shunts, and ETV intention to treat. In B are shown representative CT images for cases of normal brain volume at 24 months for female shunt (a) and female ETV (b), and growth failure at 24 months for male shunt (c) and ETV (d). Brain volume (and normalized percentage) is given on each image shown. In C are shown normalized brain volumes, age adjusted and plotted as z-scores, for volumes measured before surgery, and 6, 12, and 24 months following surgery. Cases with substantial growth are indicated with X’s, and filled colored circles indicate substantial volume loss. The horizontal line at z=-1 indicates the threshold of normal age-adjusted volume (above) and volume less than normal for age (below).

### Comparison of volume and developmental outcomes

At 24 months, those with normal brain volume had higher median scores in all BSID-3 domains and in all BSID-3 changes from baseline (**Table 3**). Moderate (Spearman rho >0.4) and statistically significant correlations were seen between 24-month brain volume (raw and normalized) and all BSID-3 scores and BSID-3 changes from baseline (**Table 4**). Only low correlations (Spearman rho <0.4) were seen with CSF volume and all developmental outcomes, however.

**Table 3.** Developmental outcome results for brain growth*.

|  | Difference (95% confidence interval) for those with *normal volume at 24 months* (N=16/90) compared to not | Difference (95% confidence interval) for those achieving *substantial brain growth* (N=23/89) compared to not | Difference (95% confidence interval) for those having *substantial brain volume loss* (N=19/89) compared to not |
| --- | --- | --- | --- |
| BSID Scaled Scores at 24 months |  |  |  |
| BSID Cognitive | 4 (2, 4) | 0 (-1, 2) | 0 (-1, 1) |
| BSID Gross Motor | 6 (2, 7) | 0 (-1, 0) | 0 (0, 0) |
| BSID Fine Motor | 4 (2, 6) | 0 (-1, 2) | -0 (-2, 2) |
| Change in BSID Scaled Scores at 24 months from baseline |  |  |  |
| BSID Cognitive, change from baseline | 4 (2, 6) | 1 (0, 3) | 0 (-1, 3) |
| BSID Gross Motor, change from baseline | 4 (2, 7) | 0 (-1, 1) | 1 (-1, 3) |
| BSID Fine Motor, change from baseline | 4 (2, 5) | 2 (0, 3) | 0 (-2, 2) |

\* Hodges-Lehmann estimates and 95% confidence intervals were used for differences in medians.

**Table 4.** Correlation of 24 month developmental outcome with brain and fluid volume*.

|  | Raw brain volume,<br>cm<sup>3</sup> | Normalized brain<br>volume, z-score | CSF volume, cm<sup>3</sup> |
| --- | --- | --- | --- |
| BSID Scaled Scores at 24 months |  |  |  |
| BSID Cognitive | 0.63<br>P<0.001 | 0.64<br>P<0.001 | -0.29<br>P=0.006 |
| BSID Gross Motor | 0.52<br>P<0.001 | 0.52<br>P<0.001 | -0.31<br>P=0.003 |
| BSID Fine Motor | 0.58<br>P<0.001 | 0.61<br>P<0.001 | -0.23<br>P=0.03 |
| Change in BSID Scaled Scores at 24 months from baseline |  |  |  |
| BSID Cognitive,<br>change from<br>baseline | 0.48<br>P<0.001 | 0.48<br>P<0.001 | -0.06<br>P=0.61 |
| BSID Gross Motor,<br>change from<br>baseline | 0.45<br>P<0.001 | 0.46<br>P<0.001 | -0.25<br>P=0.02 |
| BSID Fine Motor,<br>change from<br>baseline | 0.55<br>P<0.001 | 0.56<br>P<0.001 | -0.08<br>P=0.44 |

\* Spearman correlations and p-values shown. Data available for 88 patients.

## DISCUSSION

Our longitudinal prospective results have revealed the novel finding of a particular temporal pattern in brain growth following treatment of infant post-infectious hydrocephalus in sub-Saharan Africa. Specifically, we found that important gains are made in normalized brain volume in the first year after surgical treatment,^14^ but these are largely lost by 2 years as brain growth stagnates in the second 12 months. As a result, on average, these infants (who started out with 15% having normal brain volume at baseline) lost virtually all the brain volume gains they made in the first year (where 50% had normal brain volume) by the end of the second year (where only 18% had normal brain volume). The raw brain volume results indicate that the decline in normalized brain volumes between 12 and 24 months did not reflect a true absolute loss of volume. Rather, brain volumes stagnated between 12 and 24 months, rather than increasing, as would normally be expected for infants of that age. The important clinical implication of this is emphasized by the consistent positive correlation we observed between brain volume and developmental outcome. Our findings raise key questions that could have relevance to the treatment and outcome of infant hydrocephalus, especially in regard to this and other causes of post-inflammatory hydrocephalus (such as post-hemorrhagic hydrocephalus of prematurity).^22,23^

First, what is the mechanism of brain growth early after treatment of infant hydrocephalus? Hydrocephalus causes cortical mantle compression, decreased cerebral blood flow, demyelination, and axonal degeneration.^24,25^ Postnatal human brain growth normally results more from synaptogenesis and myelination, than from neuronal cellular proliferation.^26^ The majority of neurogenesis is complete at term, while gliogenesis peaks toward the end of gestation and continues through the first year of life. It is likely, but not proven, that early increase in brain volume following hydrocephalus treatment in the first year reflects tissue decompression and re-expansion of cortical mantle, along with increased cerebral blood flow and blood volume. Our study was only able to assess brain volume by CT imaging, an unavoidable limitation in most sub-Saharan Africa settings, which limited our ability to examine brain structural content in more detail. MRI imaging would, for instance, enable us to segment and quantify white from grey matter growth (for which we are developing normative curves), and quantify myelination during the critical second year of life. Regardless of the underlying cellular compartments responsible, however, the correlation of such increases in total brain volume with developmental outcome emphasize the importance of this growth.

Second, why did brain growth stagnate in the second year after treatment? By 2 to 3 years of age, the human brain is expected to achieve 80 - 90% % of its adult volume,^15^ which adds particular importance to our assessment of brain volume at 2 years after treatment. In our cohort of infants, beyond the first year following surgery, the potential for brain growth appeared to have declined. The initial brain injury from infection and the inflammatory response in the early weeks of life could impact brain development differently at different stages due to temporal variations in the dominant mechanisms of brain growth. The effects of early brain injury could be projected over time with differential effects on future phases of brain development. The fact that there was no difference in these outcomes between treatment groups, that raw brain volume initially increased and then stabilized (rather than declined), and that loss of normalized brain volume in the second year was the same regardless of the extent of brain growth in the first year, all suggest that brain growth and development might have been impacted more by the initial insult than by hydrocephalus or its mode of treatment. That is, it was the brain’s intrinsic growth potential that may have determined outcome at 2 years.

There are, however, other potential explanations, as well. In the specific case of post-infectious hydrocephalus in Uganda, it is also possible that ongoing, and heretofore unrecognized, indolent brain infection could further hinder growth. Whether children with post-infectious hydrocephalus and apparently culture-negative ventricular CSF may harbour an occult, indolent bacterial or viral (e.g. cytomegalovirus) infection at the time of initial hydrocephalus treatment is a current area of investigation.^27^ Certainly, the prevalence of unrelated diseases known to impact childhood survival in our patient population, such as malaria and malnutrition, may also hinder brain growth and development, especially when gauged against norms for infants in high-income countries. The future availability of normative brain growth curves reflective of Ugandan children would improve the comparison against existing norms.

Third, should hydrocephalus treatment target brain volume growth rather than CSF reduction? This would represent a paradigm shift in our thinking about hydrocephalus, but our data suggest that it is larger brain volume – independent of CSF volume – that is the major determinant of outcome. Regardless of treatment, at 2 years, we continued to see significant positive correlations between brain volume and cognitive outcome that was not seen with CSF volume. For the time being, it remains technically challenging to obtain accurate brain volume measurements in a rapid-enough fashion to guide clinical treatment decisions in most sub-Saharan African settings. We anticipate, however, that with the emergence of available inexpensive low-field MRI technologies^28,29^ and continuing development of more automated machine learning segmentation protocols,^30^ brain volume could become part of standard hydrocephalus assessment and follow-up in the way that ventricle size is currently.

### Limitations of the study

This study only addressed post-infectious hydrocephalus and this represents one of the most severely affected forms of infant hydrocephalus, with very low brain volumes and poor developmental performance at baseline. Although post-infectious hydrocephalus is the most common cause of infant hydrocephalus in sub-Saharan Africa - and likely globally, as well - we cannot comment on the developmental and brain growth outcomes for other types of infant hydrocephalus. It is possible that in those with greater structural brain integrity at baseline, the temporal pattern of post-treatment brain growth might be different. The study was conducted at a high-volume center with a great deal of experience in treating infant hydrocephalus; these results might not apply to low-volume or less experienced centers. Our assessment of developmental outcome was limited to a portion of the BSID-3 and further limited by the generally poor-function of this group of infants. More robust testing at an older age could provide different results and, indeed, we intend to follow this cohort longer-term to establish 5-year outcome results. All of our results will, of course, need to be replicated in other infant cohorts to see if similar findings are obtained. The National Institutes of Health has, in fact, funded another large randomized trial that will address these issues in infants with hydrocephalus in North America (NCT04177914) using detailed magnetic resonance imaging and developmental outcomes.

## Conclusion

Even after successful surgical treatment of infant post-infectious hydrocephalus in sub-Saharan Africa, early post-treatment brain growth in the first year stagnates in the second year. Our findings underscore the importance of primary infection prevention and mitigation strategies along with optimizing the child’s environment to maximize brain growth potential and minimize the long-term effects of the initial brain insult.

## Data Availability

Data from this study will be made available to qualified researchers by contacting the authors.

## Abbreviations

BSID: Bayley Scales of Infant Development
CCHU: CURE Children’s Hospital of Uganda
CSF: cerebrospinal fluid
CT: computed tomography
ETV/CPC: endoscopic third ventriculostomy plus choroid plexus cauterization
VPS: ventriculoperitoneal shunt

## APPENDIX

### Steering committee

Benjamin C Warf (Harvard University, USA), Steven J Schiff (Pennsylvania State University, USA), Abhaya V Kulkarni (University of Toronto, Canada)

### Investigators

Edith Mbabazi (CURE Children’s Hospital of Uganda, Uganda), John Mugamba (CURE Children’s Hospital of Uganda, Uganda), Peter Ssenyonga (CURE Children’s Hospital of Uganda, Uganda), Ruth Donnelly (The Hospital for Sick Children, Canada), Jody Levenbach (The Hospital for Sick Children, Canada), Vishal Monga (Pennsylvania State University, USA), Mallory Peterson (Pennsylvania State University, USA), Venkateswararao Cherukuri (Pennsylvania State University, USA)

### Data Safety Monitoring Board

Jay Riva-Cambrin (Chair) (University of Calgary, Canada), Michael Scott (Harvard University, USA), Graham Fieggen (University of Cape Town, South Africa), Julius Kiwanuka (Mbarara University of Science & Technology, Uganda), Francis Bajunirwe (Mbarara University of Science & Technology, Uganda)

### Research assistants

Esther Nalule, Julie Johnson, John Kimbugwe, Brian Nsubuga Kaaya

The steering committee designed the trial and analysis plan, wrote the manuscript, and takes responsibility for protocol adherence and study results. All steering committee members and investigators approved the final manuscript. The independent Data Safety Monitoring Board provided oversight of the study conduct. None of the authors have any conflicts of interest to report.

